# Direct access physiotherapy to help manage patients with musculoskeletal disorders in an emergency department: results of a randomized controlled trial

**DOI:** 10.1101/2020.10.28.20221531

**Authors:** Rose Gagnon, Kadija Perreault, Simon Berthelot, Eveline Matifat, François Desmeules, Bertrand Achou, Marie-Christine Laroche, Catherine Van Neste, Stéphane Tremblay, Jean Leblond, Luc J. Hébert

## Abstract

**Context:** In several countries, physiotherapists (PT) have been integrated within emergency departments (EDs) to help manage patients with musculoskeletal disorders (MSKDs). Still, research on the effects of such initiatives is scarce.

**Objectives:** To evaluate the effects of direct access PT on MSKD patients consulting the ED in terms of clinical outcomes and use of health care resources.

**Design, Setting, Participants:** Randomized controlled trial, academic ED in Quebec City (Canada), participants 18-80 years presenting with a minor MSKD.

**Intervention:** Direct access PT at the ED

**Control:** Emergency Physicians lead management (EP).

**Main Outcome Measure:** Clinical outcomes (pain, interference of pain on function) and use of resources (ED return visit, interventions, diagnostic tests, consultations) were compared between groups at ED discharge and after 1 and 3 months using two-way ANOVAs, log-linear analysis and χ^2^ tests.

**Results:** Seventy-eight patients suffering from MSKDs were included (40.2 ± 17.6 years old; 44% women). Participants in the PT group (n=40) had statistically lower levels of pain and pain interference at 1- and 3-months. They were recommended fewer imaging tests (38% vs. 78%; p<.0001) and prescription medication (43% vs. 67%; p=.030) at ED discharge, had used less prescription medication (32% vs. 72%; p=.002) and had revisited significantly less often the ED (0% vs. 21%; p=.007) at 1-month than those in the EP group (n=38). At 3 months, the PT group had used less over-the-counter medication (19% vs. 43%; p=.034).

**Conclusion:** Patients presenting with a MSKD to the ED with direct access to a PT had better clinical outcomes and used less services and resources than those in the usual care group after ED discharge and up to 3 months after discharge. The results of this study support the implementation of such models of care for the management of this population.

**Trial Registration:** This trial is registered at the US National Institutes of Health (ClinicalTrials.gov) #NCT04009369

**Ethical approval:** This trial was approved by the Research Ethics Committee of the CHU de Québec - Université Laval #MP-20-2019-4307

## Introduction

Musculoskeletal disorders (MSKDs) are highly prevalent and are often associated with pain, stiffness, loss of joint mobility, deformity and/or physical limitations. ^1^ MSKDs are one of the major drivers of increases in years lived with disability, low back pain alone being the worldwide leading cause of this worrisome finding. ^2^ Direct costs of MSKDs are rapidly increasing, including costs for consultations with healthcare professionals, imaging tests and medication. ^3,4^ Indirect costs, such as loss of productivity, are however considered to be the largest contributing factor to MSKDs expenditures. ^4^ For low back pain, inflation-adjusted societal costs were estimated at 26.40 billion USD in the United Kingdom and 81.24 billion USD in the United States in 2015. ^5^

The emergency department (ED) remains one of the most common setting patients turn to when presenting a MSKD. ^6^ For example, in Australia, up to 25% of ED presentations are for MSKDs. ^7^ In Canada, 9% of patients presenting to the ED suffer from low back pain. ^8^ Numerous reasons have been put forward to explain why persons go to the ED for such conditions. In a 2007 American survey, more than 40% of patients believed that their primary care provider was incapable of correctly managing MSKDs. ^9^ Other reasons include lack of access to a general practitioner (office closed, absence of a general practitioner, no health care insurance plan), functional loss (impaired walking), feeling that current pain was different from other episodes, desire for quick pain relief or additional investigation including imaging tests, as well as advice of others to consult the ED. ^10^

Therefore, MSKDs may contribute to ED overcrowding which is a major problem in health systems worldwide. ^11^ This phenomenon has numerous consequences such as prolonged lengths of stay, increased rates of patients leaving the ED without being seen, increased medical errors, increased mortality among ambulatory and non-ambulatory patients and decreased patient satisfaction. ^12–16^ In order to reduce ED overcrowding and associated long wait times, physiotherapists (PT) have been integrated in EDs in several countries to manage patients with MSKDs directly after triage by the ED nurse, in models often referred to as direct access or advanced practice physiotherapy in the ED, depending on the context. ^17,18^ Access to physiotherapy services in a timely manner for MSKDs has been associated with a decrease in psychological symptoms, decreased risks of developing persistent pain, decreased costs and utilization of the health care services. ^19–24^ Country-wide initiatives to integrate PTs in EDs in Australia and the UK have been found to reduce wait times, length of stay, the prescription of unnecessary consultations and useless diagnostic tests. ^7,17,25,26^ Some studies show promising results regarding diagnostic agreement between PTs and ED physicians or other specialists ^27^ and high patient satisfaction regarding PT care in the ED. ^28^ However, to the best of our knowledge, to date, only three randomized controlled trials assessing the effects of the integration of PTs in EDs have been performed. ^29–31^ Furthermore, very few studies have measured the effects over time on the clinical course of patients and the use of services and resources.

Therefore, to address this shortcoming in the current state of knowledge, the objectives of our project were to compare the effects of direct access physiotherapy to usual care provided by an emergency physician (EP) for ED patients presenting with a MSKD on the clinical course of patients (pain and pain interference) and the use of resources at ED discharge and at 1 and 3 months.

## Methods

### Study design

A randomized controlled trial was carried out at the CHU de Québec - Université Laval within the *Centre Hospitalier de l’Université Laval* (CHUL), an academic ED located in Quebec City (Canada). Two groups of participants were recruited over a 24-week period between September 2018 and March 2019: one group of participants having direct access to a PT (Gr-PT) and a control group (Gr-CTL) managed according to usual practice by the EP. To control for the types of MSKD between groups, participants were stratified according to the area of the body affected (lumbar spine and lower limb or thoracic spine, cervical spine and upper limb). Block randomization was used to assign them to either group. The feasibility of the randomization method was validated during a three-day observation period before data collection. The randomization sequence was generated by the principal investigator (LJH) and group allocation was unveiled by RG who opened a sealed envelope at the ED after triage by the ED nurse and patient enrollment. Follow-ups at 1 and 3 months were made either by mail or over-the-phone. The study was approved by the Research Ethics Committee of the CHU de Québec - Université Laval and registered at the US National Institutes of Health #NCT04009369.

### Study population

Eligible patients presented to the ED with a potential peripheral or vertebral minor MSKD, traumatic or not, and were given a triage score of 3 (urgent), 4 (less urgent) or 5 (non urgent) according to the Canadian Emergency Department Triage and Acuity Scale classification. ^32^ Further inclusion criteria were: 1) being aged between 18 and 80 years, 2) having the ability to legally consent to participate, 3) understanding French to complete the study questionnaires either orally or in writing, and 4) being a beneficiary of the provincial health insurance plan (*Régie de l’assurance maladie du Québec*). Patients were excluded if they presented with a major MSKD requiring emergent care (e.g. open fracture, dislocation, open wound), a red flag (e.g. progressive neurological disorder, infectious symptoms), a concomitant unstable clinical condition (e.g. pulmonary, cardiac, digestive and/or psychiatric), or if they were currently hospitalized or lived in a long-term care facility.

### Study recruitment

Potential participants were identified based on information collected by the triage nurse and included within the electronic information system used at the CHUL to register patients. After identification in the system, the research coordinator (RG) went to the waiting room to speak with the patient, verify eligibility, describe the research project, answer questions, and obtain a written informed consent. Each participant was then allocated to either group via the previously stated randomization procedure. All refusals were recorded and documented.

### Study interventions

Participants in the PT group were initially assessed by a PT following nurse triage. After obtaining a brief history of the injury and of clinical signs and symptoms, the PT performed a brief physical examination of each patient. Interventions were then recommended based on the clinical analysis and physiotherapy diagnosis, including advice, technical aids, imaging, prescribed or over-the-counter medication, and consults with other healthcare professionals.

Immediately after each consultation, the PT filled a standardized form containing a summary of the initial assessment, including diagnosis, and the recommended clinical management. He also completed his usual clinical note in the patient’s medical record. The form and a copy of the note were then added to the ED consultation request. In the context of this innovative study, the CHUL internal policy was to have every patient presenting to the ED seen by the ED physician prior to discharge. The EP was free to use the PT’s recommendations or not but was encouraged to consult and discuss with the PT, if judged relevant. As for participants in the control group, they received usual care consisting in the ED physician assessment followed by their choice of interventions that were documented in the patient’s file.

### Measures

Our primary outcomes were pain interference on function and pain intensity which were measured before ED management and at 1- and 3-months post ED visit. They were measured using the *Brief Pain Inventory* (BPI) and the *Numeric Pain Rating Scale* (NPRS), respectively, both tools having been recognized as being reliable, valid and responsive. ^33–37^ They were administered using paper format at the ED. Secondary variables related to utilization of services and resources at ED discharge were documented from the above-mentioned forms and patient’s paper and electronic record. They included the following: types of interventions recommended (advice and education, exercises, technical aid, hospitalization, medication), healthcare professionals consulted, and imaging tests recommended. The BPI, NPRS, ED return visits, hospitalization rate, number and types of consultations, imaging tests and medication used were collected at 1- and 3-months following ED discharge using a self-reported online questionnaire sent to participants via email or through structured telephone interviews, according to patient preference and availability.

### Data analysis and sample size

Descriptive statistics were used to characterize participants. Two-sample Kolmogorov-Smirnov tests and χ^2^ tests were used to compare characteristics of participants between groups at baseline. We used non-parametric repeated-measures analyses of variance (two-way ANOVAs) for longitudinal data (R software, 3.6.1; package nparLD, 2.1; proc f1.LD.f1) to measure differences between groups over time for BPI and NPRS scores (2 groups x 3 times). These analyses are more robust than parametric and semiparametric procedures as they can accept a change in distribution over time, are unaffected by extreme values and can be used with smaller samples or ordinal scales. ^38^ Furthermore, nparLD ANOVAs are designed so that there is no need to impute values when patients are lost to follow-up. Hence, per-protocol analyses were conducted. ^38,39^

Multiway frequency analysis were performed to compare secondary variables between groups across time points (SPSS 25, proc hiloglinear). ^40^ χ^2^ tests were used to compare data between groups at each time point individually (producing exact p-values). The α criterion was set at .05 for all statistical analyses.

An a priori sample size was based on the minimum clinically important difference (MCID) of the BPI estimated to be 1.00. ^41,42^ Using G*Power 3.1.9.2, we estimated a required sample-size of 45 participants per group based on α=0.05, effect size=0.66, power [1-β]=0.80, standard deviation 1 (SD1)=1.43, SD2=1.59 BPI points, MCID=1.00 BPI points, loss to follow-up=20%.

## Results

### Participants

Overall, 579 participants were assessed for eligibility between September 2018 and March 2019 and 78 were recruited (Figure 1). Ten participants declined to participate because they wanted to be exclusively managed by the EP. Two participants did not receive the allocated intervention in the EP group because they left the ED before seeing the EP, but they still participated in the 1- and 3-months follow-ups. Fifteen participants were lost to follow-up at 1 month (follow-up rate: 80.86% [CTL group: 84.22%; PT group: 77.5%]) but 4 of them were successfully contacted at 3 months. Sixteen participants were lost to follow-up at 3 months (follow-up rate: 79.47% [CTL group: 80.0%; PT group: 78.95%]). No adverse event was reported.

**Figure 1.**
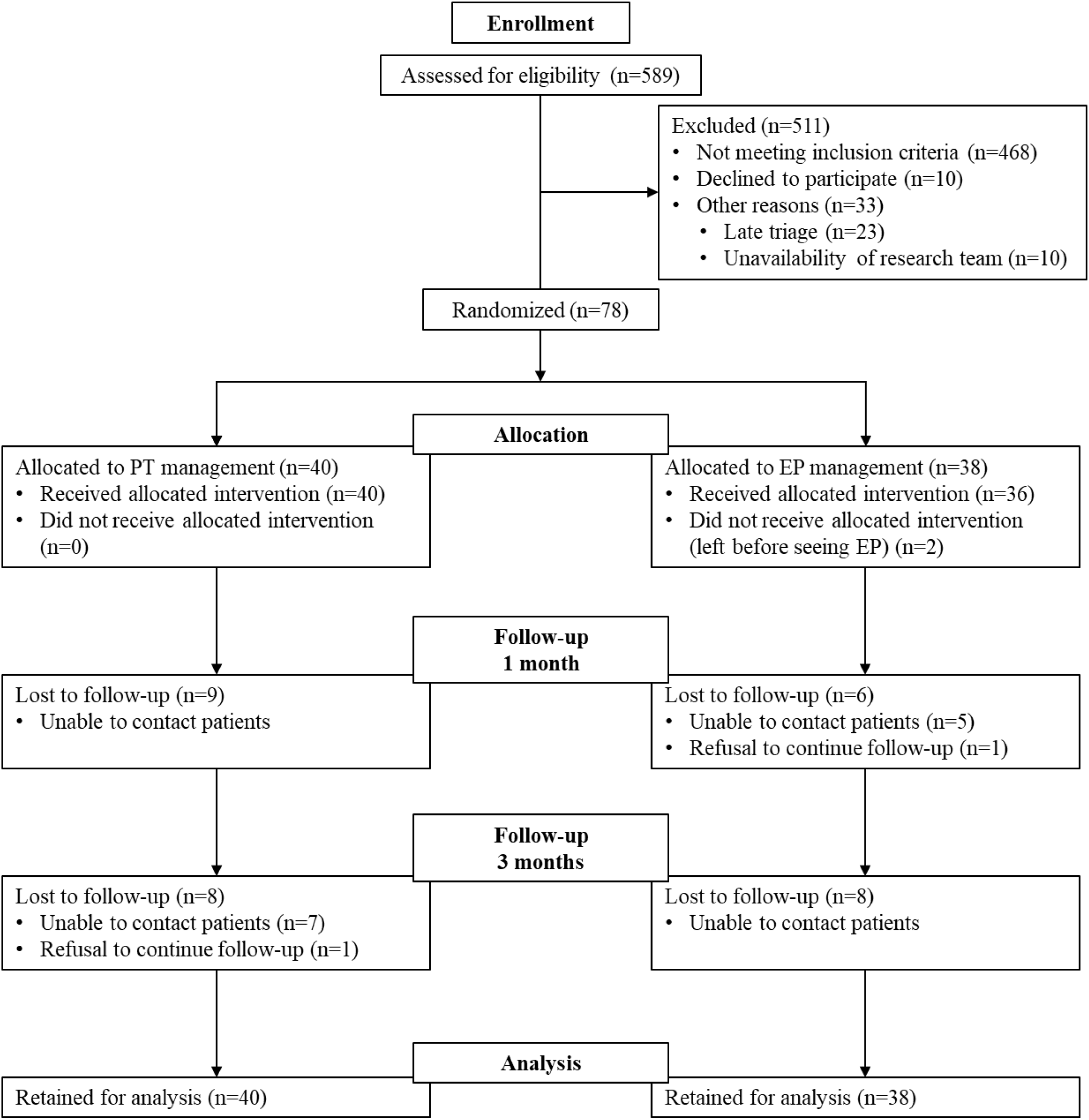
CONSORT Flow Diagram of participants through the trial.

Regarding baseline characteristics, there were no statistically significant differences between groups for all variables, except for age (p=.007) and gender (p=.043) (Table 1). There were more women in the PT group and their mean age was significantly lower. Hence, we used these characteristics as covariates in our analysis.

**Table 1.** Baseline characteristics of study participants (n=78)

| Characteristics | CTL (n=38) | PT (n=40) |
| --- | --- | --- |
| Age (yrs), mean (SD)<sup>1</sup> | 44.08 (17.3) | 36.55 (17.3) |
| Gender, n males (%)<sup>2</sup> | 26 (68.4) | 18 (45.0) |
| Triage category in ED, n (%) |  |  |
| Urgent (P3) | 16 (42.1) | 16 (40.0) |
| Semi-urgent (P4) | 21 (55.3) | 24 (60.0) |
| Non-urgent (P5) | 1 (2.6) | 0 (0) |
| Living arrangements, n (%) |  |  |
| Alone | 8 (21.1) | 10 (25.0) |
| With family/Spouse | 30 (78.9) | 30 (75.0) |
| Occupation, n (%) |  |  |
| Medical leave due to present complaint | 2 (5.3) | 3 (7.5) |
| Medical leave for other reason | 3 (7.9) | 0 (0) |
| Working Full time | 19 (50.0) | 18 (45.0) |
| Working Part time | 4 (10.5) | 2 (5.0) |
| Student | 3 (7.9) | 10 (25.0) |
| Other | 7 (18.4) | 7 (17.5) |
| Loss of wages since onset, yes n (%) | 3 (7.9) | 3 (7.5) |
| Personal income, n (%) |  |  |
| 1\$ - 49 999\$ | 24 (63.2) | 25 (62.5) |
| 50 000\$ - 99 999\$ | 11 (28.9) | 11 (27.5) |
| ≥ 100 000\$ | 1 (2.6) | 0 (0) |
| Highest level of education completed, n (%) |  |  |
| Primary or High School | 14 (36.8) | 5 (12.5) |
| College or University | 24 (63.2) | 34 (85.0) |
| Time since onset of symptoms, n weeks (%) |  |  |
| 0-2 | 32 (84.2) | 30 (75.0) |
| 3-12 | 1 (2.6) | 4 (10.0) |
| > 12 | 4 (10.5) | 4 (10.0) |
| Don't know | 1 (2.6) | 1 (2.5) |
| Other health conditions, yes n (%) | 18 (47.4) | 13 (32.5) |
| Mode of arrival in the ED, n (%) |  |  |
| Ambulance | 7 (18.4) | 6 (15.0) |
| Private or public transport | 29 (76.3) | 28 (70.0) |
| Foot | 1 (2.6) | 6 (15.0) |
| Other | 0 (0) | 0 (0) |
| Registered to a family physician, n (%) | 30 (78.9) | 33 (82.5) |
| Pain catastrophizing, mean (SD) | 22.4 (11.8) § | 18.3 (13.2) † |

1: p<.05, 2: p<.01, §: n=35, †: n=39, CTL: Control group, PT: PT group, Yrs: years, ED: Emergency department

### Effect of direct access PT on pain intensity and pain interference

Our analysis showed significant group effect, time effect and group x time effect on the BPI and NPRS scores (Table 2, Figures 2a.-2b.). Both groups showed significant improvements over time (NPRS: p<.0001, BPI: p<.0001; Table 2), but participants in the PT group had a greater improvement at the 1- and 3-month follow-ups compared to the CTL group (NPRS: p=.0005, BPI: p=.048; Table 2). Both groups achieved minimal clinically important differences for the NPRS (MCID: 1.3 points) and BPI scores (MCID 1.0 points) at 1 month, but scores in the PT group were significantly lower than those in the CTL group (Table 2, Figures 3a.-3b.).

**Table 2.**
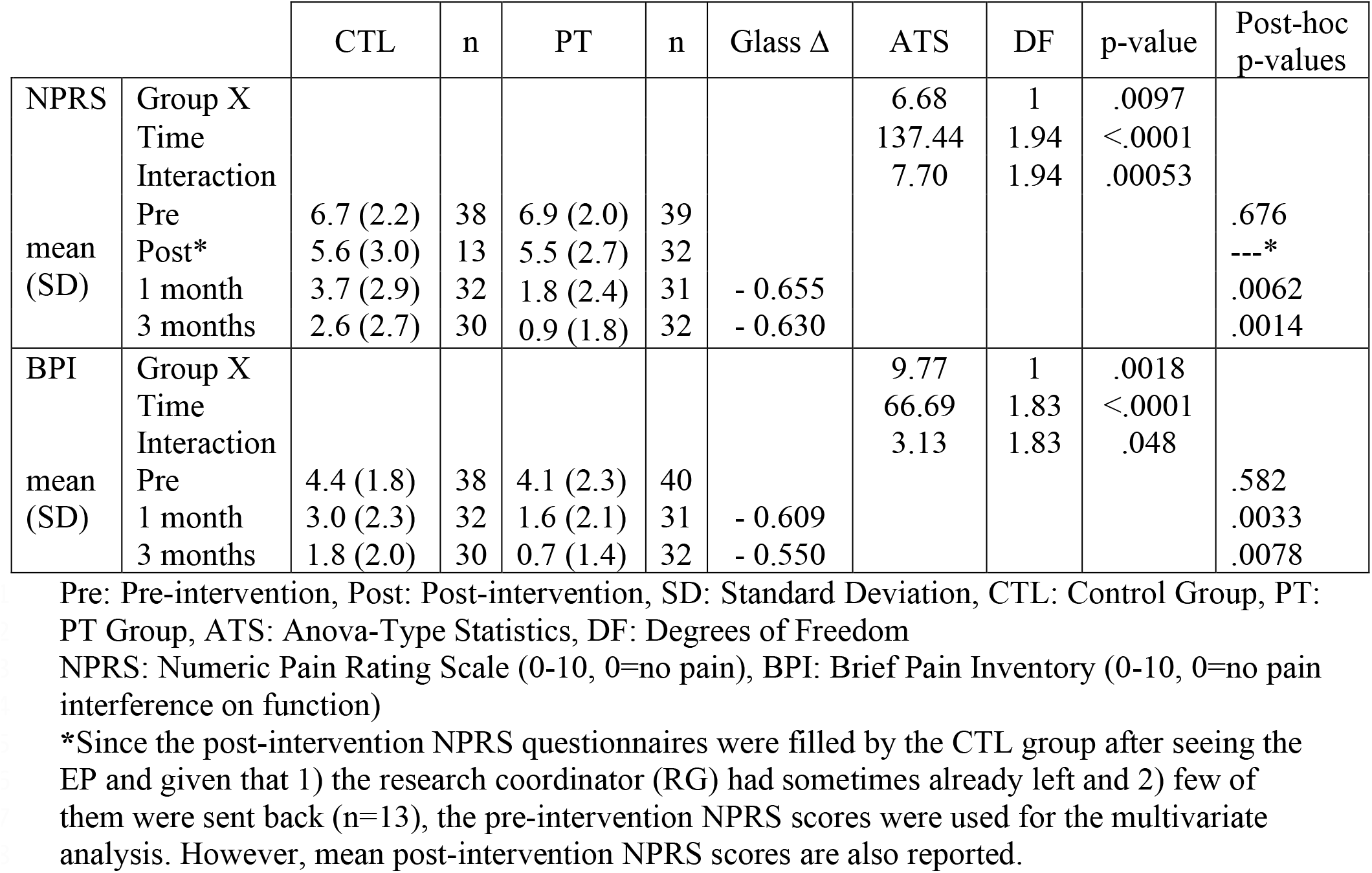
Pain intensity and pain interference at baseline, 1- and 3-month follow-ups.

| | | CTL | n | PT | n | Glass $\Delta$ | ATS | DF | p-value | Post-hoc<br>p-values |
| --- | --- | --- | --- | --- | --- | --- | --- | --- | --- | --- |
| NPRS | Group X |  |  |  |  |  | 6.68 | 1 | .0097 |  |
|  | Time |  |  |  |  |  | 137.44 | 1.94 | <.0001 |  |
|  | Interaction |  |  |  |  |  | 7.70 | 1.94 | .00053 |  |
|  | Pre | 6.7 (2.2) | 38 | 6.9 (2.0) | 39 |  |  |  |  | .676 |
|  | Post* | 5.6 (3.0) | 13 | 5.5 (2.7) | 32 |  |  |  |  | ---* |
| mean<br>(SD) | 1 month | 3.7 (2.9) | 32 | 1.8 (2.4) | 31 | - 0.655 |  |  |  | .0062 |
|  | 3 months | 2.6 (2.7) | 30 | 0.9 (1.8) | 32 | - 0.630 |  |  |  | .0014 |
| BPI | Group X |  |  |  |  |  | 9.77 | 1 | .0018 |  |
|  | Time |  |  |  |  |  | 66.69 | 1.83 | <.0001 |  |
|  | Interaction |  |  |  |  |  | 3.13 | 1.83 | .048 |  |
|  | Pre | 4.4 (1.8) | 38 | 4.1 (2.3) | 40 |  |  |  |  | .582 |
|  | 1 month | 3.0 (2.3) | 32 | 1.6 (2.1) | 31 | - 0.609 |  |  |  | .0033 |
| mean<br>(SD) | 3 months | 1.8 (2.0) | 30 | 0.7 (1.4) | 32 | - 0.550 |  |  |  | .0078 |

Pre: Pre-intervention, Post: Post-intervention, SD: Standard Deviation, CTL: Control Group, PT:
PT Group, ATS: Anova-Type Statistics, DF: Degrees of Freedom
NPRS: Numeric Pain Rating Scale (0-10, 0=no pain), BPI: Brief Pain Inventory (0-10, 0=no pain interference on function)
\*Since the post-intervention NPRS questionnaires were filled by the CTL group after seeing the EP and given that 1) the research coordinator (RG) had sometimes already left and 2) few of them were sent back (n=13), the pre-intervention NPRS scores were used for the multivariate analysis. However, mean post-intervention NPRS scores are also reported.

**Figure 2.**
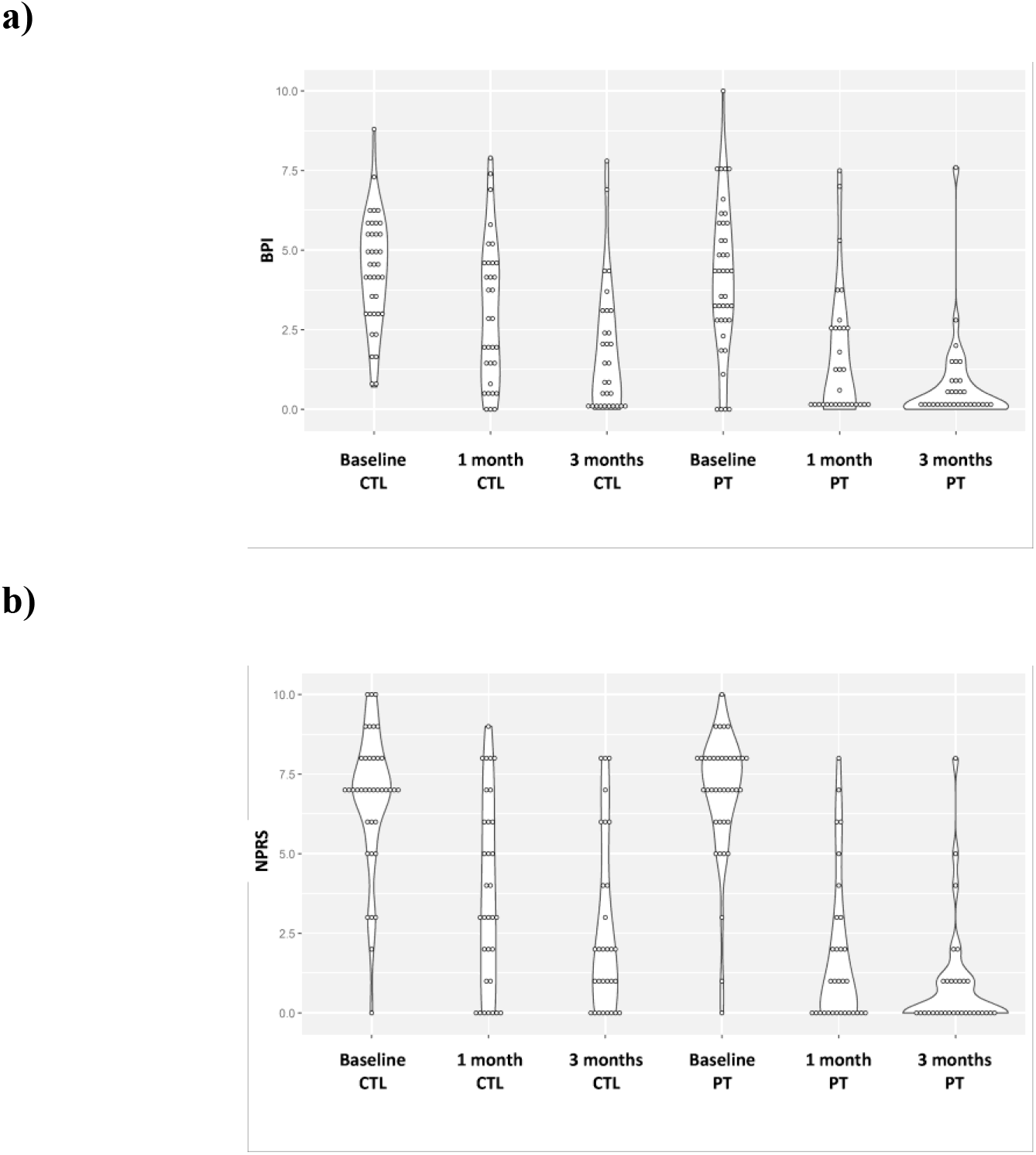
Violin plot of the scores for each group at baseline, 1 and 3 months. a)BPI scores and b) NPRS scores. CTL: Control group PT: PT group BPI: Brief Pain Inventory (0-10, 0=no pain interference on function) NPRS: Numeric Pain Rating Scale (0-10, 0=no pain)

**Figure 3.**
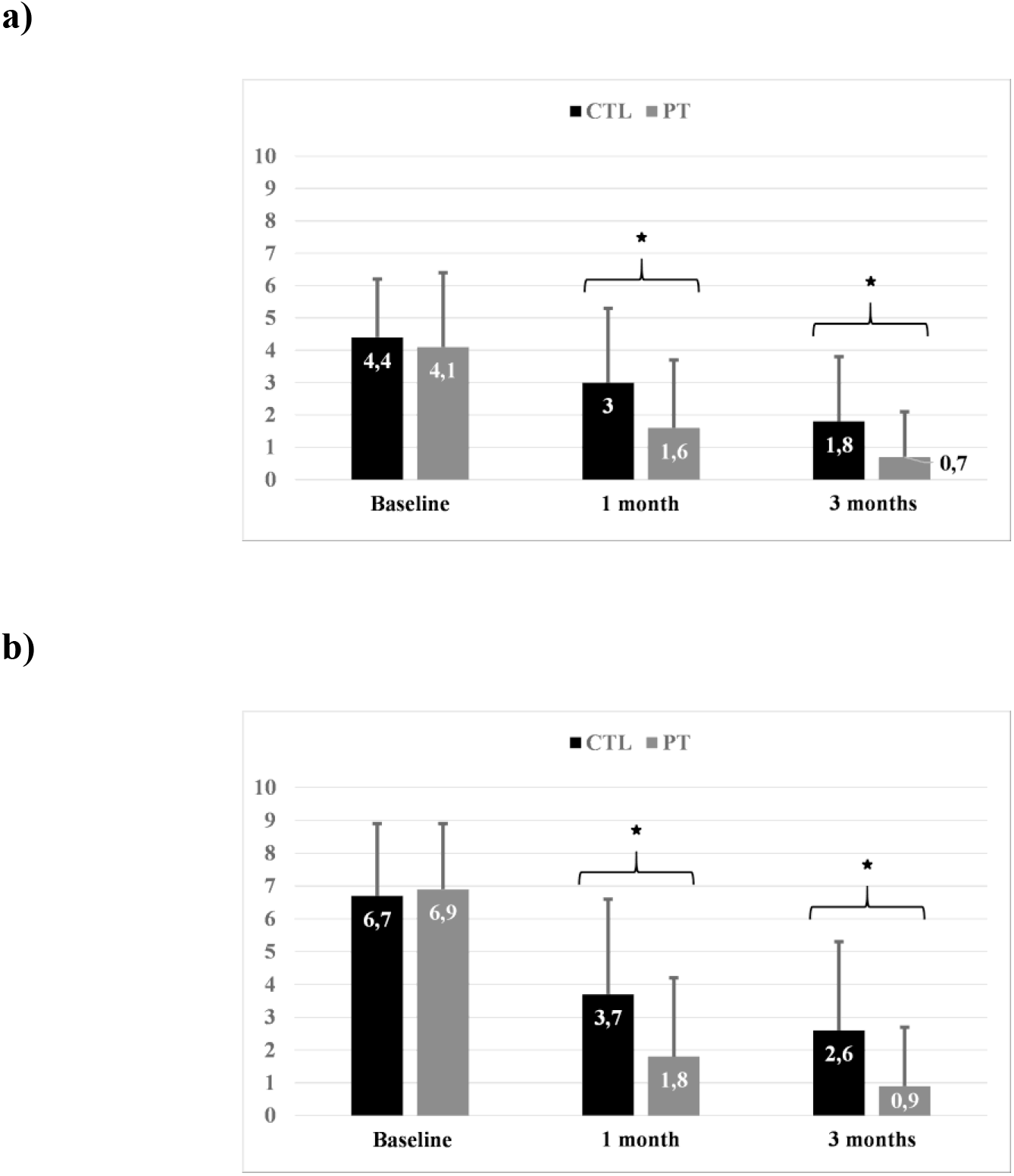
Mean scores for each group at baseline, 1 and 3 months. a) BPI scores and b) NPRS scores. CTL: Control group PT: PT group *: p<.05 BPI: Brief Pain Inventory (0-10, 0=no pain interference on function) NPRS: Numeric Pain Rating Scale (0-10, 0=no pain)

Differences in scores in the PT group remained significantly lower than those in the CTL group at 3 months; mean scores of participants seen by the PT were 2.9 times lower for the NPRS and 2.6 times lower for the BPI compared to the control group at the 3-month follow-up (Table 2). To calculate the effect size, Glass’s *delta* were used since the standard deviation of both groups were statistically different.

### Comparisons of use of services and resources

We found significant differences between groups at ED discharge regarding proportions of prescription and over-the-counter medication and imaging tests (only recommended in the case of the PT but autonomously prescribed in the case of the EP). The PT recommended significantly less prescribed medication (p=.030; Figures 4a.-5) and imaging tests (p<.0001) but more over-the-counter medication (p<.0001) than the EP at the ED. After 1 month, participants in the PT group had returned significantly less often to the ED (p=.007) and had used less prescription medication (p=.002), including opioids (p=.043), than participants in the control group (Figures 4b.-5). There were no differences in the use of over-the-counter medication (p=.102), other professionals consulted (p=.269), and imaging tests administered (p=.477). At 3 months, participants in the PT group used less over-the-counter medication than participants in the EP group (p=.034), but there were no significant differences in ED return visits (p=.484), prescription medication (p=.234), imaging tests used (p=.150) and consultations with another professional (p=.503) (Figures 4c.-5). No differences were found in hospitalization rates between groups at all time points (at discharge p=.474; 1-month p=.738; 3-month no hospitalization in both groups). As seen on the Figure 5, overall, at all times, the proportion of use of services and resources between groups was higher in the ED group compared to the PT groups for all services and resources except for the over-the-counter medication (OTCM) and the imaging tests (IT).

**Figure 4.**
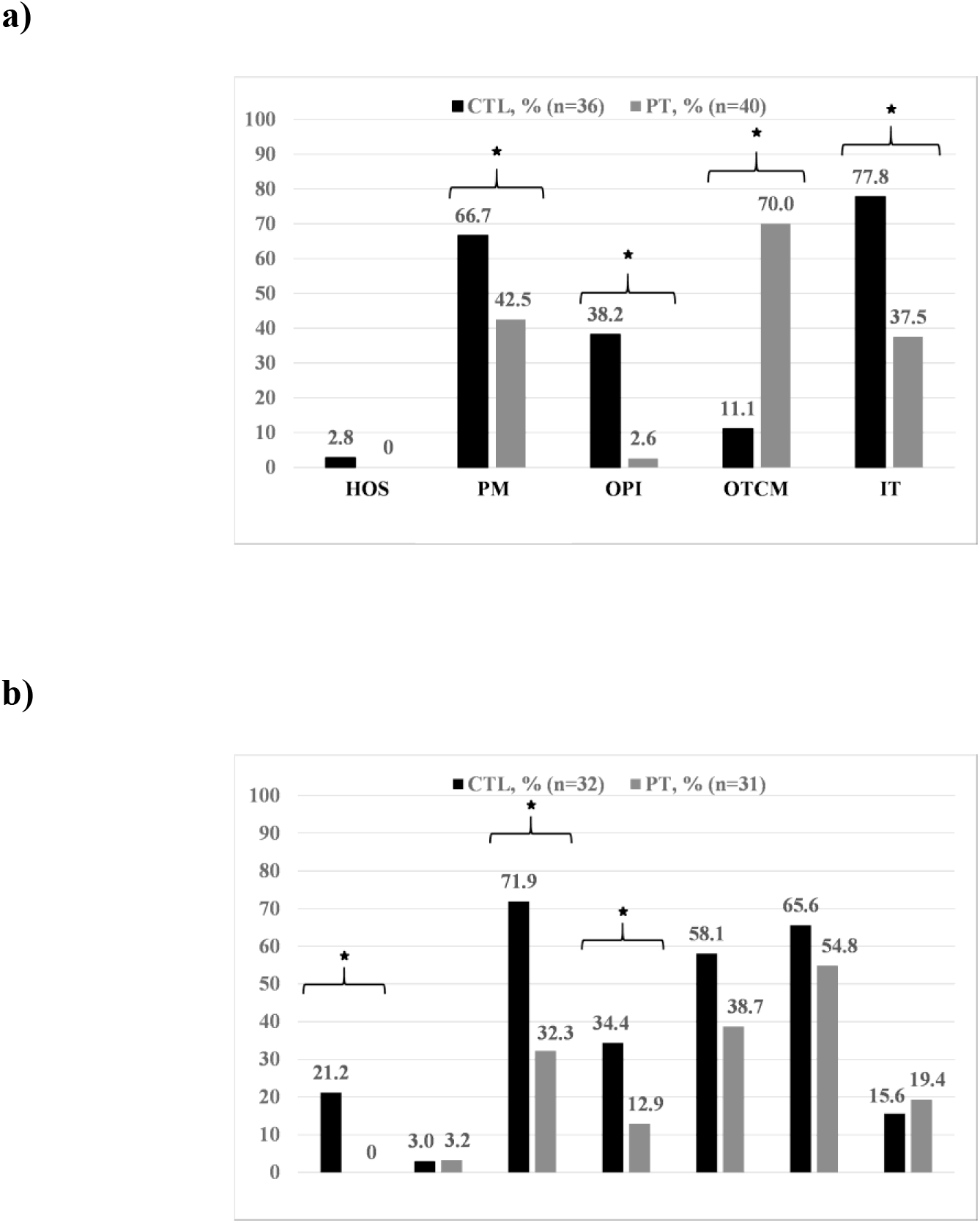
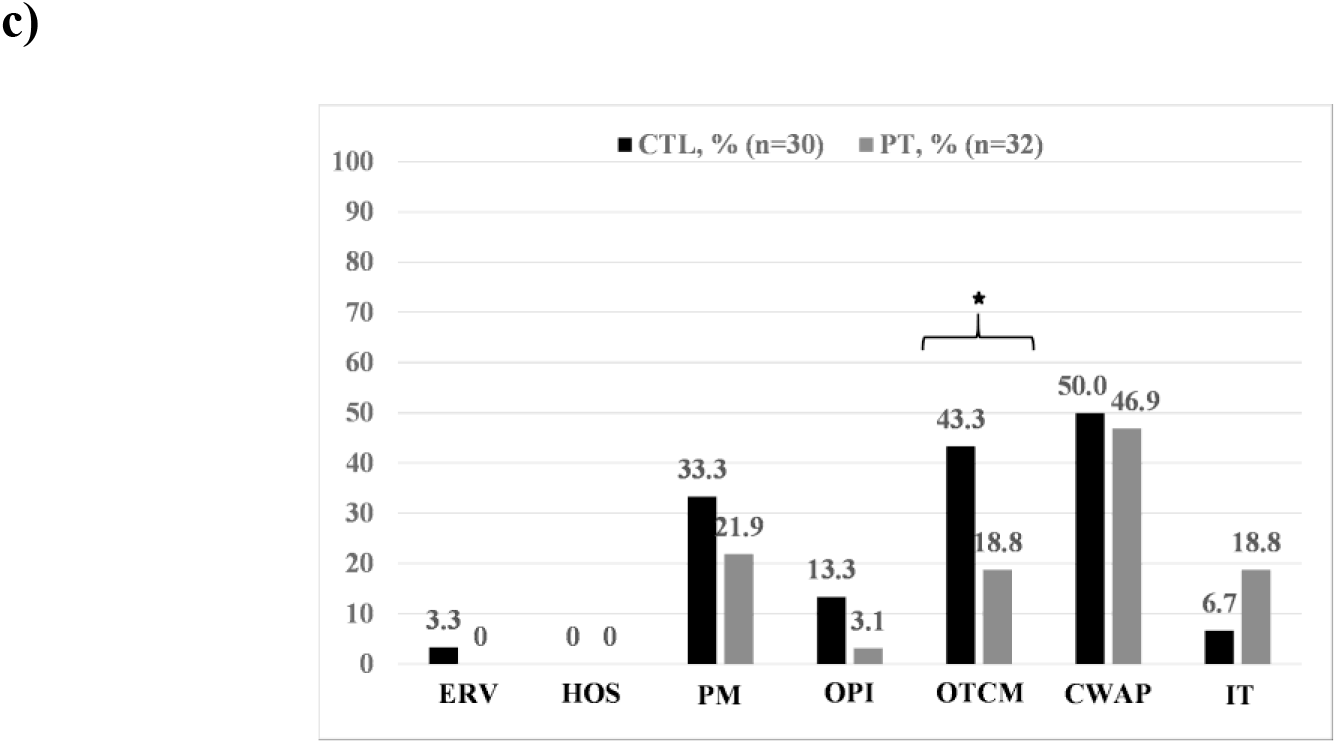
Proportion of services and resources recommended by each provider a) During ED visit, b) After 1 month and c) Between the 1-and 3-month follow-ups. CTL: Control group PT: PT group *: p<.05 ERV: ED return visits HOS: Hospitalization PM: Prescription medication OPI: Opioids OTCM: Over-the-counter medication CWAP: Consultation with another professional IT: Imaging tests

**Figure 5.**
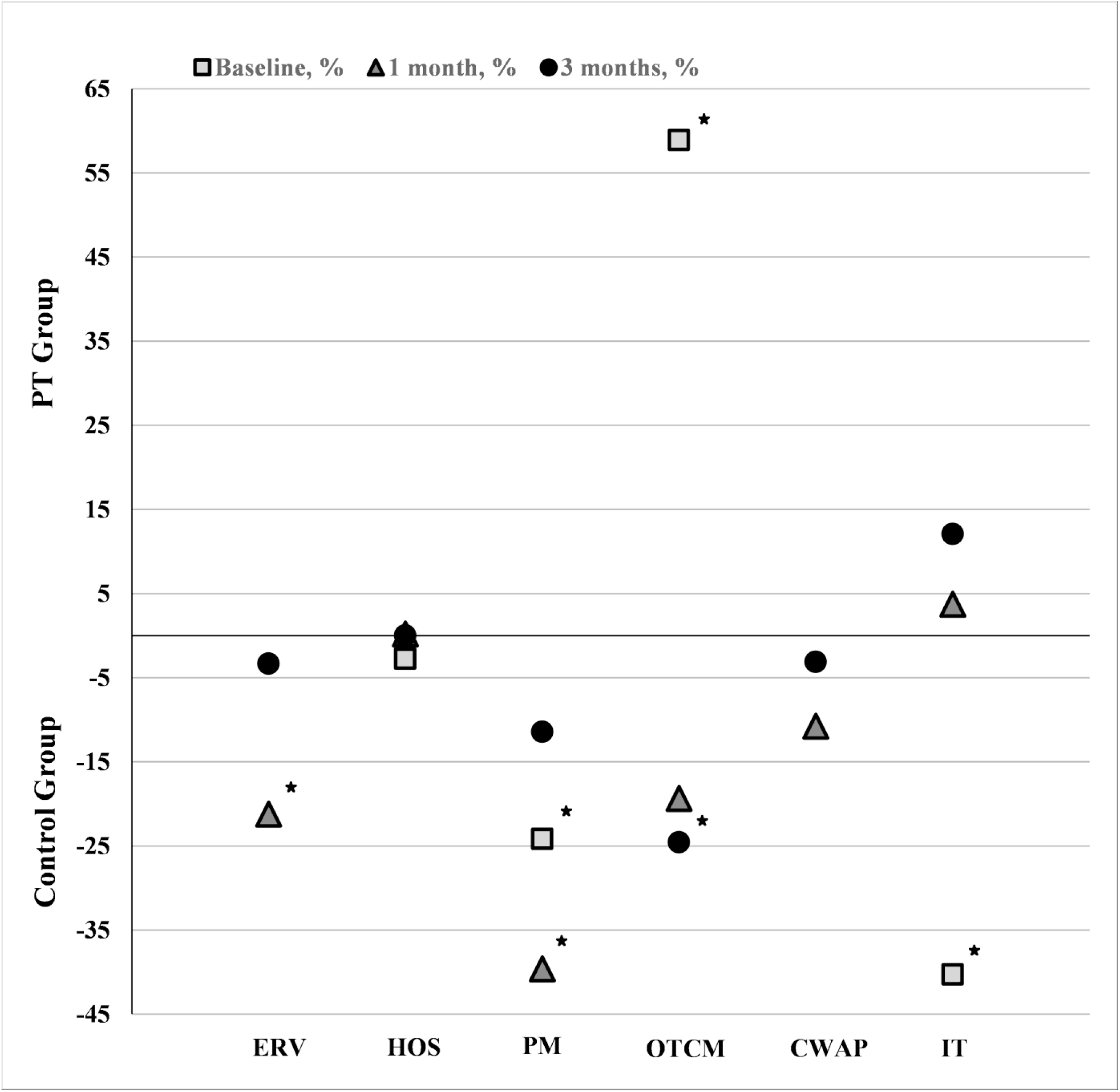
Proportion of use of services and resources between groups at baseline (■), 1 (▲) and 3 months (•). Values represent the difference between proportions in the PT group and proportions in the EP group at each time point. *: p<.05 ERV: ED return visits HOS: Hospitalization PM: Prescription medication OTCM: Over-the-counter medication CWAP: Consultation with another professional IT: Imaging tests

## Discussion

The aim of our study was to compare the effects of direct access PT for patients presenting to an ED for a MSKD to usual management provided by an EP on the clinical course of participants (pain and pain interference) and use of services and resources after ED discharge and at 1 and 3 months post-discharge.

In this study, participants in the PT group presented significantly lower pain intensity and pain interference than those in the EP group at 1 and 3 months. Significant differences in scores were found between the PT and the EP group (main effect of group) and between our measures at each time point (main effect of time): scores in the PT group were statistically lower than those in the EP group and both groups showed significant improvement over time. These differences between our groups persisted over time (interaction effect), helping to confirm that the changes in scores observed over time were significantly different depending on group allocation. Indeed, mean pain intensity level in the PT group dropped by 6.0 points between baseline and the 3-months follow-up and mean pain interference on function dropped by 3.4 points. This later finding largely exceeds clinically important differences (1.3 NPRS points; 1.00 BPI points), which represents important clinical benefits. ^34,41^ Comparatively, NPRS scores dropped by 4.1 points and BPI scores by 2.6 points in the CTL group. These differences in scores could be partially explained by the quality of the reinsurance and education given by the PT. ^43^ Other studies show mixed results for pain outcomes. In a study by Lau et al., patients seen by the PT expressed significantly less pain at discharge from the ED and within one week of discharge, but these differences were no longer significant at the 1-month follow-up. ^29^ Other studies on direct access PT or advanced practice in the ED report no significant differences in pain levels at baseline and up to six months after ED visit. ^30,44–46^ More broadly, studies concerning early access to PT in various clinical settings also suggest mixed results, patients with early access presenting either a significant decrease or no change at all in their pain level after six months. ^20,21,26^ However, as reported by Kilner et al., most of these studies are of poor methodological quality and considered as low-level evidence. ^47^ To our knowledge, this is the first study presenting data on pain interference for patients managed by a PT in the ED.

Furthermore, use of several services and resources was significantly lower for participants in the PT group compared to those in the EP group at discharge, 1 and 3 months. At discharge, participants in the EP (control) group had been prescribed about 40% more diagnostic imaging tests than what was recommended by the PT. These results are consistent with others found in the literature. ^44,48^ In recent clinical guidelines, imaging is discouraged for patients presenting with MSKDs unless a serious pathology is suspected or if imaging is likely to change management. ^49,50^ According to Baker et al., approximately 40% of imaging referrals for patients presenting with a non-traumatic MSKD to the ED is inconsistent with guideline recommendations. ^51^ Moreover, when looking at the PT’s assessment, Décary et al. found high inter-rater agreement for common knee disorders between the diagnosis made by the PT using only a musculoskeletal examination and the physician’s diagnosis made using both musculoskeletal examination and imaging. The musculoskeletal examination performed by the PT has also been found to be of high diagnostic validity. ^27^ Thus, it is possible that the differences observed in the PT’s recommendation for imaging may be due in part to the more comprehensive physical examination performed or greater adherence to clinical practice guidelines. No adverse outcomes were reported in both groups. There were no differences between groups for use of imaging tests at 1 and 3 months, suggesting that direct access PT management was appropriate an at least equivalent to that of the EP.

At ED discharge, participants in the PT group were also recommended on average 25% less often prescription medication but around 60% more over-the-counter medication than those in the control group. Furthermore, participants in the PT group had used 40% less prescription medication at 1 month, including opioids, and 25% less over-the-counter medication at 3 months compared to the EP group. In agreement with certain clinical guidelines, acetaminophen, anti-inflammatory drugs or opioids should not be recommended to patients presenting with a MSKD as their efficacy is questionable and they tend to be associated with poorer outcomes. ^49,50^ Providers should prioritize modalities such as advice to stay active, individualized patient education and supervised exercise. ^49,50^ As for ED return visits, no participant in the PT group had returned to the ED in the first 3 months after their ED visit. Comparatively, 21% of participants seen by the EP at 1 month had returned to the ED. To our knowledge, this is the first time that an effect on ED return visits and medication use is measured in a study comparing the effects of management of MSKD by a PT or an EP in ED. The differences in ED return visits rates might be partially explained by the different training and approaches used by the two care providers. As mentioned above, current guidelines recommend providing education or information to encourage self-management and to inform and reassure patients about their condition. ^49^ Patients should also be offered individualised education in addition to usual care. ^50^ For example, pain neuroscience education for MSKDs has been found to reduce pain and improve patient knowledge of pain, improve function, enhance movement and minimize healthcare utilization. ^52^ It has been shown that PTs have higher levels of knowledge in managing MSKDs than medical students, physician interns, residents and all physician specialists except for orthopaedists ^53^. Participants in the PT group may have then felt better equipped and empowered to manage their condition which may have reduced perceived need for further services and resources over time.

## Limitations

We found significant differences in age and gender between our groups at baseline, the PT group having younger participants and more women. Such differences in allocation might have been caused by the stratification used when randomizing participants as we stratified only according to the area of the body affected and not by gender or age. Women have been found to report higher pain intensity and disability than men for the same condition, to seek more medical care, to cope less efficiently with pain and to be managed differently, also receiving more prescription medication and at higher doses than men. ^54,55^ Since there were more women in the PT group than the EP group, the positive effects of the PT intervention over time might have been under-estimated. However, we used age and gender as covariates in our analysis and hence controlled for this limitation. Also, the groups may have differed for other baseline characteristics that were only present in few participants, thus preventing us from verifying whether a statistical difference was present.

Sample size in our study is relatively small. We must then be reasonably careful with the generalization of the results, although sufficient statistical power was obtained. Moreover, we only had a 20% loss to follow-up. It is possible that quality of the data may have been compromised by recall bias and lack of completeness of the notes written by each of the professionals involved. Another limitation, implicitly imposed from an ethical point of view, is that we did not include a group receiving no treatment at all, hence preventing us from comparing the evolution of our participants with the expected natural healing process overtime. Also, for feasibility reasons, longer-term follow-up of participants was not carried out.

## Conclusion

The results of this study suggest that direct access PT in the ED for patients presenting with a MSKD is associated with greater improvements in pain intensity and pain interference and less use of several services and resources, such as ED return visits, imaging tests, prescription and over-the-counter medication. Further multicentric trials are needed to confirm these findings. Moreover, future studies should include an economic analysis to ascertain if PT management in a direct access setting in an ED is cost-effective.

## Supporting information

CONSORT checklist

## Data Availability

The data that support the findings of this study are available from the corresponding author, LJH, upon reasonable request.

## Acknowledgements

We would like to thank the following persons for their contribution to project implementation:

All project participants, Antony Barabé PT, physiotherapist at the Centre Hospitalier de l’Université Laval (CHUL), Myriam Mallet, research professional, Jasmine Martineau who was the senior manager of health care professional services when the project was initiated, and Mathieu Blanchet, MD, FRCPC, head of the CHUL ED department during the duration of the study.

## References

1. Agel J, Akesson K, Amadio PC, Anderson M, Rasker JJ. The burden of musculoskeletal conditions at the start of the new millennium. 2003 [cited 2019 Jun 28];Available from: https://research.utwente.nl/en/publications/the-burden-of-musculoskeletal-conditions-at-the-start-of-the-new-

2. GBD 2015 Disease and Injury Incidence and Prevalence Collaborators. Global, regional, and national incidence, prevalence, and years lived with disability for 310 diseases and injuries, 1990-2015: a systematic analysis for the Global Burden of Disease Study 2015. Lancet Lond Engl 2016;388(10053):1545–602.

3. Lin C-WC, Haas M, Maher CG, Machado LAC, van Tulder MW. Cost-effectiveness of general practice care for low back pain: a systematic review. Eur Spine J 2011;20(7):1012–23.

4. Bornhöft L, Thorn J, Svensson M, Nordeman L, Eggertsen R, Larsson MEH. More cost-effective management of patients with musculoskeletal disorders in primary care after direct triaging to physiotherapists for initial assessment compared to initial general practitioner assessment. BMC Musculoskelet Disord [Internet] 2019 [cited 2019 May 8];20(1). Available from: https://bmcmusculoskeletdisord.biomedcentral.com/articles/10.1186/s12891-019-2553-9

5. Hartvigsen J, Hancock MJ, Kongsted A, et al. What low back pain is and why we need to pay attention. The Lancet 2018;391(10137):2356–67.

6. Jordan KP, Kadam UT, Hayward R, Porcheret M, Young C, Croft P. Annual consultation prevalence of regional musculoskeletal problems in primary care: an observational study. BMC Musculoskelet Disord 2010;11(1):144.

7. Bird S, Thompson C, Williams KE. Primary contact physiotherapy services reduce waiting and treatment times for patients presenting with musculoskeletal conditions in Australian emergency departments: an observational study. J Physiother 2016;62(4):209–14.

8. Institut canadien d’information sur la santé. SNISA - Nombre de visites au service d’urgence et durée de séjour par province et territoire, 2015-2016. 2016;

9. Gaieski DF, Mehta S, Hollander JE, Shofer F, Bernstein J. Low-severity Musculoskeletal Complaints Evaluated in the Emergency Department. Clin Orthop 2008;466(8):1987–95.

10. Stafford V, Greenhalgh S, Davidson I. Why do patients with Simple Mechanical Back Pain seek Urgent Care? Physiotherapy 2014;100(1):66–72.

11. Di Somma S, Paladino L, Vaughan L, Lalle I, Magrini L, Magnanti M. Overcrowding in emergency department: an international issue. Intern Emerg Med 2015;10(2):171–5.

12. Epstein SK, Huckins DS, Liu SW, et al. Emergency department crowding and risk of preventable medical errors. Intern Emerg Med 2012;7(2):173–80.

13. Forero R, Man N, Ngo H, et al. Impact of the four-hour National Emergency Access Target on 30 day mortality, access block and chronic emergency department overcrowding in Australian emergency departments. Emerg Med Australas 2019;31(1):58–66.

14. Bernstein SL, Aronsky D, Duseja R, et al. The Effect of Emergency Department Crowding on Clinically Oriented Outcomes. Acad Emerg Med 2009;16(1):1–10.

15. Santos E, Cardoso D, Queirós P, Cunha M, Rodrigues M, Apóstolo J. The effects of emergency department overcrowding on admitted patient outcomes: a systematic review protocol. JBI Database Syst Rev Implement Rep 2016;14(5):96.

16. Carter EJ, Pouch SM, Larson EL. The Relationship Between Emergency Department Crowding and Patient Outcomes: A Systematic Review. J Nurs Scholarsh 2014;46(2):106–15.

17. de Gruchy A, Granger C, Gorelik A. Physical Therapists as Primary Practitioners in the Emergency Department: Six-Month Prospective Practice Analysis. Phys Ther 2015;95(9):1207–16.

18. Morris J, Vine K, Grimmer K. Evaluation of performance quality of an advanced scope physiotherapy role in a hospital emergency department. Patient Relat Outcome Meas 2015;6:191–203.

19. Foster NE, Hartvigsen J, Croft PR. Taking responsibility for the early assessment and treatment of patients with musculoskeletal pain: a review and critical analysis. Arthritis Res Ther 2012;14(1):205.

20. Wand BM, Bird C, McAuley JH, Doré CJ, MacDowell M, De Souza LH. Early Intervention for the Management of Acute Low Back Pain: A Single-Blind Randomized Controlled Trial of Biopsychosocial Education, Manual Therapy, and Exercise. Spine 2004;29(21):2350–6.

21. Nordeman L, Nilsson B, M??ller M, Gunnarsson R. Early Access to Physical Therapy Treatment for Subacute Low Back Pain in Primary Health Care: A Prospective Randomized Clinical Trial. Clin J Pain 2006;22(6):505–11.

22. Childs JD, Fritz JM, Wu SS, et al. Implications of early and guideline adherent physical therapy for low back pain on utilization and costs. BMC Health Serv Res 2015;15:150.

23. Ojha HA, Wyrsta NJ, Davenport TE, Egan WE, Gellhorn AC. Timing of Physical Therapy Initiation for Nonsurgical Management of Musculoskeletal Disorders and Effects on Patient Outcomes: A Systematic Review. J Orthop Sports Phys Ther 2016;46(2):56–70.

24. Deslauriers S, Déry J, Proulx K, et al. Effects of waiting for outpatient physiotherapy services in persons with musculoskeletal disorders: a systematic review. Disabil Rehabil 2019;0(0):1–10.

25. Heywood JW. Specialist physiotherapists in orthopaedic triage-the results of a military spinal triage clinic. J R Army Med Corps 2005;151(3):152–156.

26. Sohil P, Hao PY, Mark L. Potential impact of early physiotherapy in the emergency department for non-traumatic neck and back pain. World J Emerg Med 2017;8(2):110–5.

27. Décary S, Fallaha M, Pelletier B, et al. Diagnostic validity and triage concordance of a physiotherapist compared to physicians’ diagnoses for common knee disorders. BMC Musculoskelet Disord 2017;18(1):445.

28. Matifat E, Perreault K, Roy J-S, et al. Concordance between physiotherapists and physicians for care of patients with musculoskeletal disorders presenting to the emergency department. BMC Emerg Med 2019;19(1):67.

29. Lau PM-Y, Chow DH-K, Pope MH. Early physiotherapy intervention in an Accident and Emergency Department reduces pain and improves satisfaction for patients with acute low back pain: a randomised trial. Aust J Physiother 2008;54(4):243–9.

30. Richardson B, Shepstone L, Poland F, Mugford M, Finlayson B, Clemence N. Randomised controlled trial and cost consequences study comparing initial physiotherapy assessment and management with routine practice for selected patients in an accident and emergency department of an acute hospital. Emerg Med J EMJ 2005;22(2):87–92.

31. McClellan CM, Cramp F, Powell J, Benger JR. A randomised trial comparing the cost effectiveness of different emergency department healthcare professionals in soft tissue injury management. BMJ Open 2013;3(1):e001116.

32. Beveridge R, Clarke B, Janes L, et al. L’échelle canadienne de triage & de gravité pour les départements d’urgence Guide d’implantation. Can J Emerg Med 1999;1(3).

33. Jensen MP, McFarland CA. Increasing the reliability and validity of pain intensity measurement in chronic pain patients. Pain 1993;55(2):195–203.

34. Bijur PE, Latimer CT, Gallagher EJ. Validation of a Verbally Administered Numerical Rating Scale of Acute Pain for Use in the Emergency Department. Acad Emerg Med 2003;10(4):390–2.

35. Michener LA, Snyder AR, Leggin BG. Responsiveness of the numeric pain rating scale in patients with shoulder pain and the effect of surgical status. J Sport Rehabil 2011;20(1):115–28.

36. Mendoza T, Mayne T, Rublee D, Cleeland C. Reliability and validity of a modified Brief Pain Inventory short form in patients with osteoarthritis. Eur J Pain 2006;10(4):353–353.

37. Tan G, Jensen MP, Thornby JI, Shanti BF. Validation of the brief pain inventory for chronic nonmalignant pain. J Pain 2004;5(2):133–7.

38. Noguchi K, Gel YR, Brunner E, Konietschke F. nparLD?: An R Software Package for the Nonparametric Analysis of Longitudinal Data in Factorial Experiments. J Stat Softw [Internet] 2012 [cited 2020 Mar 11];50(12). Available from: http://www.jstatsoft.org/v50/i12/

39. Brunner E, Domhof S, Langer F. Nonparametric analysis of longitudinal data in factorial experiments. J. Wiley; 2002.

40. Tabachnick BG, Fidell LS, Ullman JB. Using multivariate statistics. Pearson Boston, MA; 2007.

41. Dworkin RH, Turk DC, Wyrwich KW, et al. Interpreting the Clinical Importance of Treatment Outcomes in Chronic Pain Clinical Trials: IMMPACT Recommendations. J Pain 2008;9(2):105–21.

42. Mease PJ, Spaeth M, Clauw DJ, et al. Estimation of minimum clinically important difference for pain in fibromyalgia. Arthritis Care Res 2011;63(6):821–6.

43. Matifat E, Méquignon M, Cunningham C, Blake C, Fennelly O, Desmeules F. Benefits of Musculoskeletal Physical Therapy in Emergency Departments: A Systematic Review. Phys Ther 2019;99(9):1150–66.

44. Schulz P, Prescott J, Shifman J, Fiore J, Holland A, Harding P. Comparing patient outcomes for care delivered by advanced musculoskeletal physiotherapists with other health professionals in the emergency department-A pilot study. Australas Emerg Nurs J AENJ 2016;19(4):198–202.

45. McClellan CM, Greenwood R, Benger JR. Effect of an extended scope physiotherapy service on patient satisfaction and the outcome of soft tissue injuries in an adult emergency department. Emerg Med J EMJ 2006;23(5):384–7.

46. Bornhöft L, Larsson ME, Nordeman L, Eggertsen R, Thorn J. Health effects of direct triaging to physiotherapists in primary care for patients with musculoskeletal disorders: a pragmatic randomized controlled trial. Ther Adv Musculoskelet Dis 2019;11:1759720×1982750.

47. Kilner E. What evidence is there that a physiotherapy service in the emergency department improves health outcomes? A systematic review. J Health Serv Res Policy 2011;16(1):51–8.

48. Sutton M, Govier A, Prince S, Morphett M. Primary-contact physiotherapists manage a minor trauma caseload in the emergency department without misdiagnoses or adverse events: an observational study. J Physiother 2015;61(2):77–80.

49. Lin I, Wiles L, Waller R, et al. What does best practice care for musculoskeletal pain look like? Eleven consistent recommendations from high-quality clinical practice guidelines: systematic review. Br J Sports Med 2020;54(2):79–86.

50. Stochkendahl MJ, Kjaer P, Hartvigsen J, et al. National Clinical Guidelines for non-surgical treatment of patients with recent onset low back pain or lumbar radiculopathy. Eur Spine J 2018;27(1):60–75.

51. Baker B, Kessler K, Kaiser B, et al. Non-traumatic musculoskeletal pain in Western Australian hospital emergency departments: A clinical audit of the prevalence, management practices and evidence-to-practice gaps. Emerg Med Australas 2019;31(6):1037–44.

52. Louw A, Zimney K, Puentedura EJ, Diener I. The efficacy of pain neuroscience education on musculoskeletal pain: A systematic review of the literature. Physiother Theory Pract 2016;32(5):332–55.

53. Childs JD, Whitman JM, Sizer PS, Pugia ML, Flynn TW, Delitto A. A description of physical therapists’ knowledge in managing musculoskeletal conditions. BMC Musculoskelet Disord 2005;6:32.

54. Fullerton EF, Doyle HH, Murphy AZ. Impact of sex on pain and opioid analgesia: a review. Curr Opin Behav Sci 2018;23:183–90.

55. Mills SEE, Nicolson KP, Smith BH. Chronic pain: a review of its epidemiology and associated factors in population-based studies. BJA Br J Anaesth 2019;123(2):e273–83.

